# Aligning Funding Models with Clinical Practice: Phases of Neonatal Care and the Link with Activity-Based Funding

**DOI:** 10.1101/2024.10.09.24314559

**Authors:** Dylan A Mordaunt

## Abstract

**Objective:** To explore the potential impact of partitioning neonatal unit funding into acute and sub-acute/non-acute care (SNAP) types, within a level 6 neonatal unit under the Independent Health and Aged Care Pricing Authority (IHACPA) National Efficient Price (NEP) Funding Model.

**Methods:** In a 12-month retrospective cohort from our neonatal unit, we simulated the effect of a care type change to SNAP. We then explored the trend in this activity type over the past 20 years.

**Results:** 341 patients with a length of stay greater than 10 days were identified from FY 2021-22. Modelling estimates that between 51 and 175 episodes could have a SNAP opportunity. When moderated, this corresponds with an uplift of between AUD $0.3 - 1.7M, based on 2023-24 value and NEP price.

**Conclusions:** Utilisation of a SNAP care-type change has the potential for a considerable uplift in funding for level 6 neonatal units, supporting service sustainability. This may be of use for other units, whilst the neonatal funding model continues to be reviewed and optimized by IHACPA. Implementation in our context, would require changes to both local and state funding management systems, as well as alterations in the Electronic Medical Record (EMR).

## Introduction

Neonatal care is highly specialized and complex. Major innovations introduced in neonatal and public health over the past 50 years have both markedly improved outcomes in newborns and significantly increased the cost of neonatal care ^1^. As these technologies have become more mature, and perinatal intervention at the limits of viability has become routine, with further advancements being made in terms of the gestational ages at which survival can occur, this has had two key effects ^2^.

Firstly, neonatal care has become more specialised in terms of workforce skill mix and health technologies, with the consequence of centralising the neonatal intensive care capacity in our jurisdiction in two centres ^3^. Secondly, funding models based on birth weight are not well aligned with the clinical and resulting operational context- for instance, units providing services to interstate patients may need to fund accommodation for patients with or without co-payment, whereas in some jurisdictions funding may be addressed through a separate accommodation scheme and not costed as part of the episode of care ^4^.

What this means is that as intervention continues to occur closer to the limits of viability, the funding models have not kept pace with clinical models, resulting in a situation where the Australian Independent Health and Aged Care Pricing Authority (IHACPA) National Efficient Price (NEP) Activity-Based Funding (ABF) model, has required specific adjustments to account for variance in outcomes for neonatal care diagnosis-related groups (DRGs), with a longer-term plan to continue to review the underlying groupings for alignment with good clinical care ^5^.

This is exacerbated in South Australia, where Neonatal Intensive Care Unit (NICU) and Special Care Baby Unit (SCBU) capacity is centralised in two units, following international trends ^6^. This centralisation limits the tacit separation of admissions into different phases, such as between neonatal intensive care (hyperacute care) through to step-down care, and the convalescent nutritional and low acuity monitoring support often provided in services with lower Clinical Services Capability Levels, interstate (sometimes referred to as “feeders and growers” or “transitional” phase).

The concept of separating out phases of the neonatal admission has long been undertaken, including the separation of different levels of care between NICUs and SCBUs ^7^. Further separation into transitional care is also well established ^8, 9^. Recent reviews of the NEP have advocated for unbundling neonatal or to remodel the neonatal Diagnosis-Related Groups (DRGs) to follow a gestationally rather than weight-orientated model of funding ^10^. This suggests that the current funding models are not aligned with good clinical practice and raises whether admissions should be separated into acute and sub-acute/non-acute (SNAP) care types, a common practice in other types of non-neonatal hospital care such as child and adult stroke care focused on functional gain.

In this study, we explored the predicted impact of introducing care-type change from acute to Australian National Subacute and Non-Acute Patient (AN-SNAP) care into a level 6 NICU and level 5 SCBU unit operation, and the potential impact that this would have on financing of the units. We simulated the impact of care type changes on ABF funding and making long-term estimates on the trends around the identified cohort.

## Methods

### Study

We undertook a simulation study utilising administrative data from patients treated in South Australia. That is, we simulated the effect of changing care-type from acute admitted care to AN-SNAP within a retrospective cohort, providing an estimate of the financial impact of care-type change within the unit. We subsequently looked over a longer period to determine the activity trends around this sub-cohort, to determine whether this activity was increasing or decreasing.

### Setting

Our context is a 50-bed level 6 NICU and associated level 5 SCBU, that sits alongside a level 6 obstetrics and perinatal service. This unit has a state-wide catchment from within both SA and the Northern Territory.

### Dataset and ethics

Data was accessed from our institutional case-mix system and patient administration system (PAS). This work was undertaken under the auspices of quality improvement activities for which institutional ethics review is exempted in our organisation. This assessment was made by the Southern Adelaide Local Health Network Research Governance Office, under the Soutb Australia Research Ethics and Governance Policy.

### Participants

A review of “qualified” admissions (admissions where the infant required medical input) was undertaken to evaluate the length of stay (LOS) for neonates, compared to the IHACPA average length of stay (ALOS) for the episode DRG. In-scope admissions included any neonatal episodes with more than 10 qualified bed days that did not have a principal diagnosis of being “born in hospital” (i.e. no comorbidities) or were admitted with a low acuity diagnosis-related group (D complexity DRG).

### Valuation

We revalued the cost of the admissions using the 2023-24 IHACPA NEP model, utilising all components of the model, including adjustments. This incorporated both uplift adjustments including outlier per diem payments and downward adjustments such as those from healthcare-acquired complications. These adjustments were separately and aggregated to create the total weighting for each scenario. The cost estimate was made by multiplying this NWAU estimate by the 2023-24 NEP value (AUD 6032).

### Analysis

To calculate the potential funding uplift, it was assumed any patient with a length of stay less than 1.5 times the IHACPA ALOS was not long enough to be considered as a potentially convertible episode. For the remaining episodes with >1.5 times the IHACPA ALOS, the National Weighted Average Unit (NWAU) opportunity for the potentially convertible episodes was modelled by the following:

1. A modelled SNAP point was calculated at either 1.5 times IHACPA ALOS or the IHACPA upper bound for the acute DRG,
2. Any potentially convertible episode with LOS above the IHACPA ALOS were split into two separate episodes of care,
3. The original episode (i.e. LOS equals modelled SNAP point) received the current inlier acute rate.

The new ‘SNAP’ episode (i.e. remaining qualified bed days above the modelled SNAP point) was funded as a SNAP episode of care (see Figure 1). The potential SNAP episode uplift was modelled using the funding parameters for Episode Type “‘4ES5’ – Maintenance, Age <= 17, LOS <= 91” ^5^.

**Figure 1.**
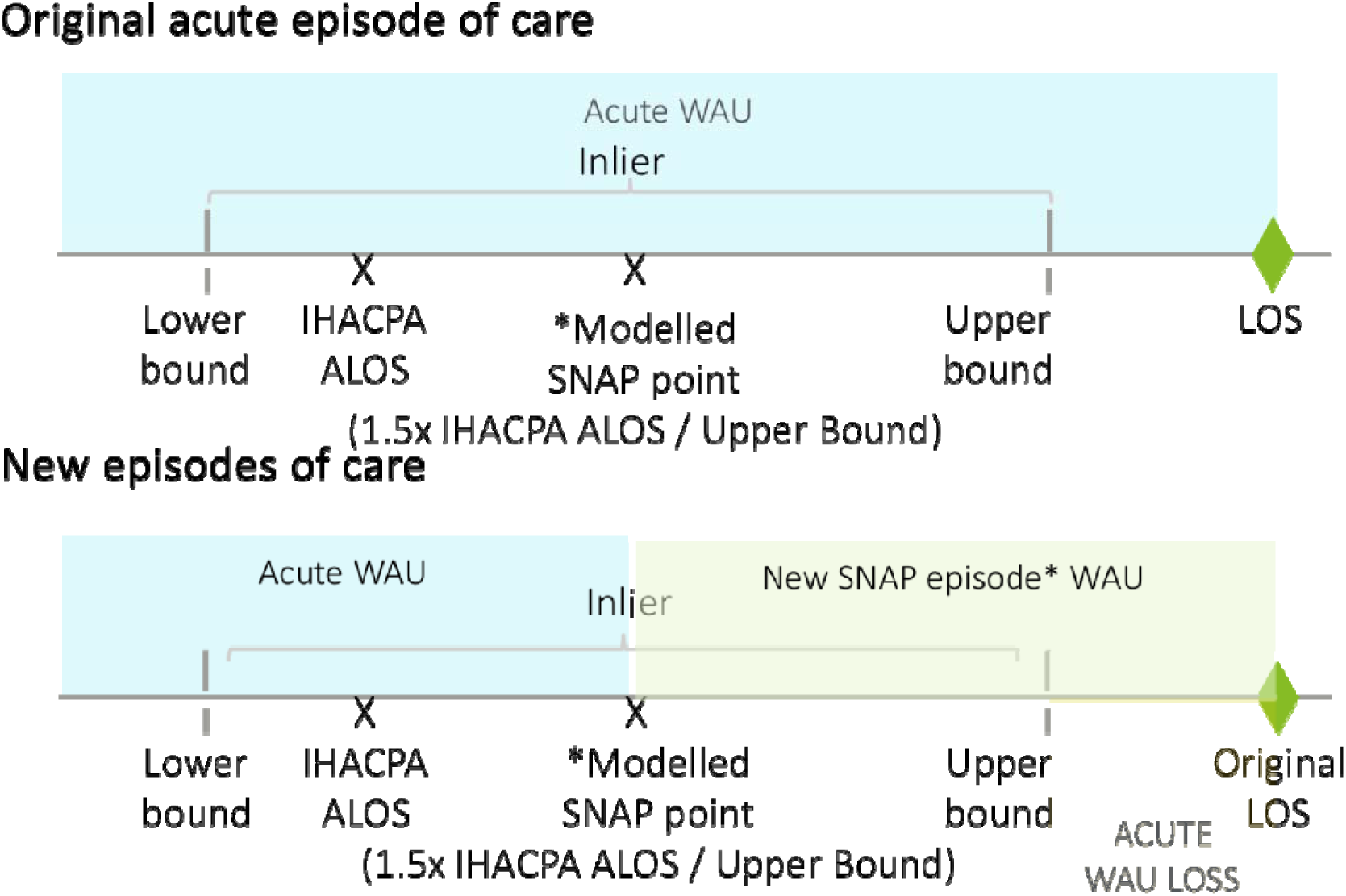
Opportunity Calculation – Qualified Neonate SNAP (Upper Estimate) Independent Health and Aged Care Pricing Authority (IHACPA), Average Length of Stay (ALOS), Length of Stay (LOS), Weighted Activity Unit (WAU).

### Long-Term Trends

To explore the long-term trends related to this opportunity, we filtered raw activity data from 2000-23 with the same inclusion and exclusion criteria and visualised these using matplotlib and Seaborn ^11, 12^.

## Results

### Baseline Characteristics

In the financial year 2021-22, there were 308 neonatal admissions deemed to be potentially convertible episodes (Figure 2), representing a total of 10,143 occupied bed days (OBDs) with an average length of stay of 32.9 days compared with the IHACPA ALOS for the same case-mix of 22.2 days. 33 of these admissions were low complexity (D complexity DRGs), and therefore excluded from further analysis. Of the remaining 308 admissions, modelling estimates that either 175 (upper estimate) or 51 (lower estimate) episodes could have a SNAP opportunity. The remaining 133 episodes were excluded due to having a LOS less than the modelled SNAP point.

**Figure 2.**
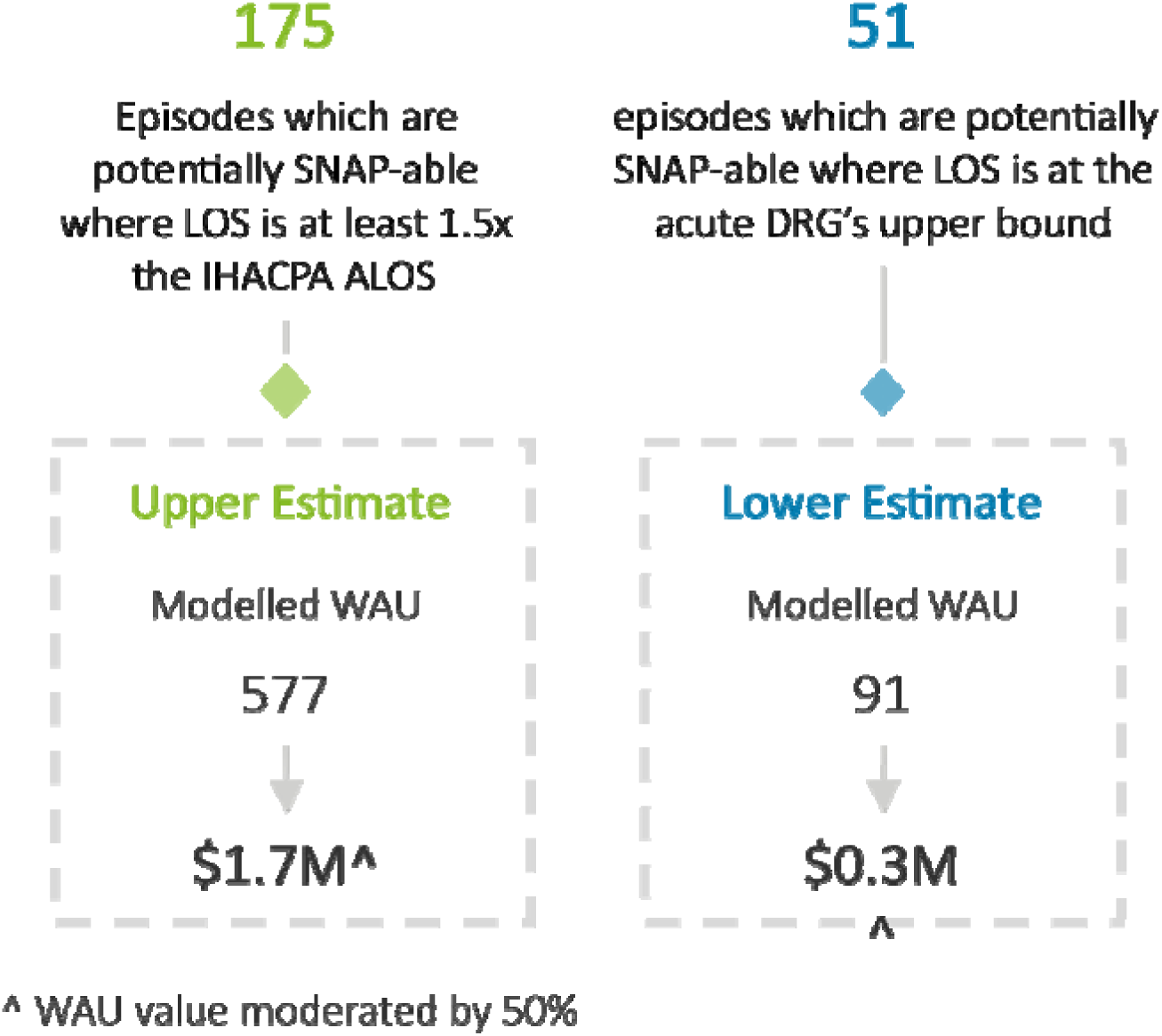
Qualified Neonate SNAP Opportunity Summary. Independent Health and Aged Care Pricing Authority (IHACPA), Average Length of Stay (ALOS), Length of Stay (LOS), Weighted Activity Unit (WAU), Diagnosis-Related Group (DRG).

**Figure 3.**
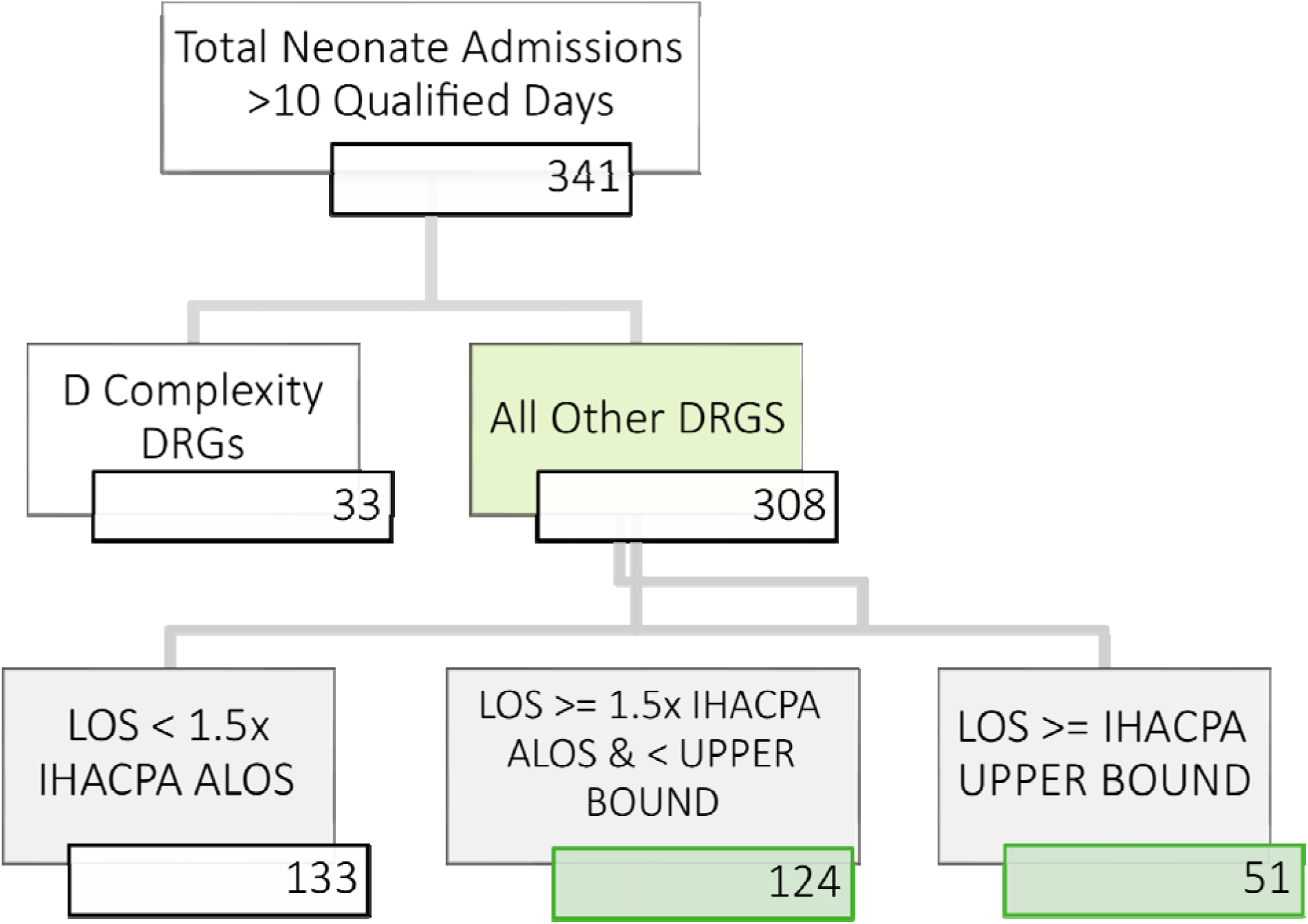
Potential Sub-Acute Neonatal Admissions Cohort Determination. Independent Health and Aged Care Pricing Authority (IHACPA), Average Length of Stay (ALOS), Length of Stay (LOS), Weighted Activity Unit (WAU), Diagnosis-Related Group (DRG).

**Figure 4.**
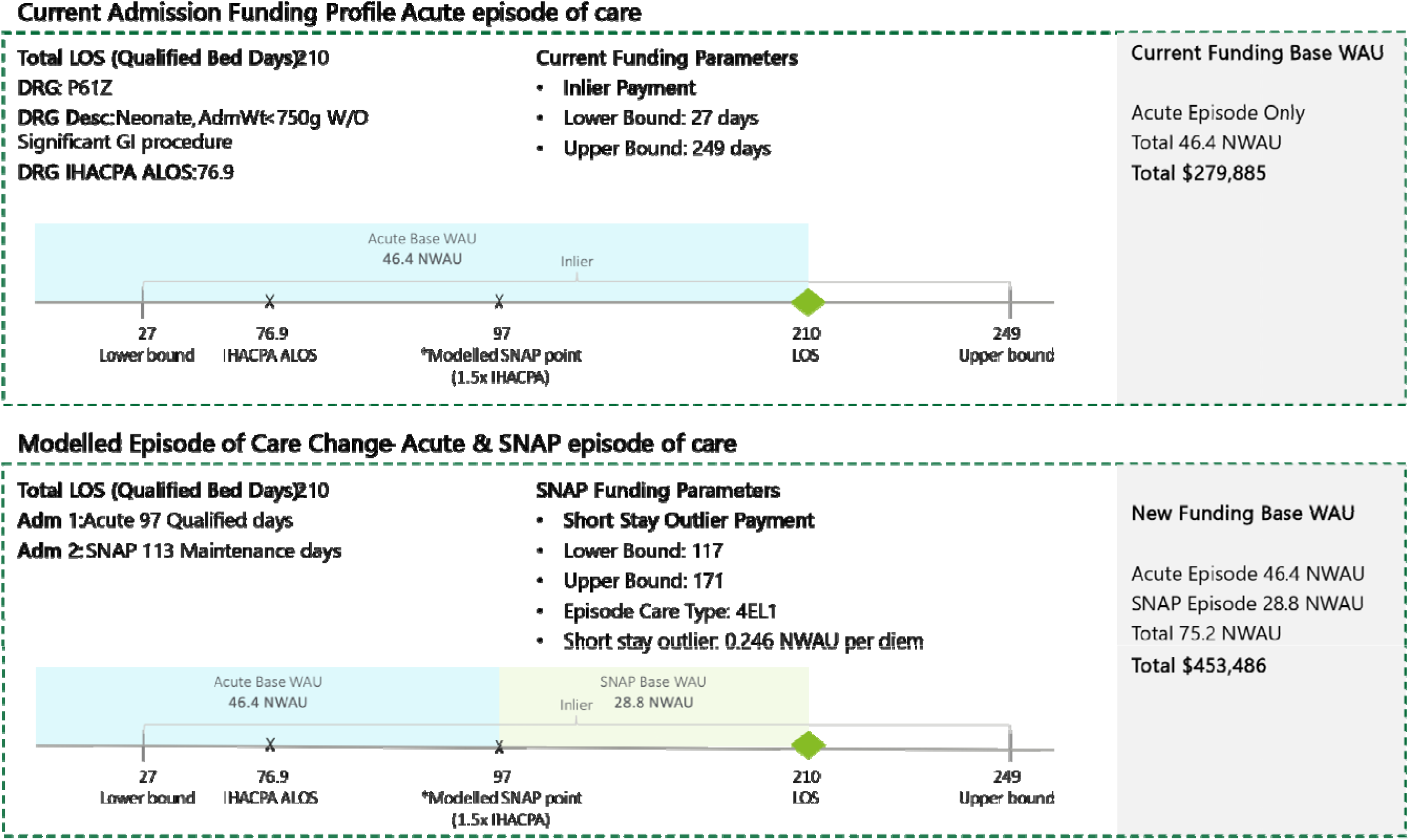
Case Study – Long-Stay Neonatal Admission - Upper Estimate. Independent Health and Aged Care Pricing Authority (IHACPA), Average Length of Stay (ALOS), Length of Stay (LOS), Weighted Activity Unit (WAU), Diagnosis-Related Group (DRG).

**Figure 5.**
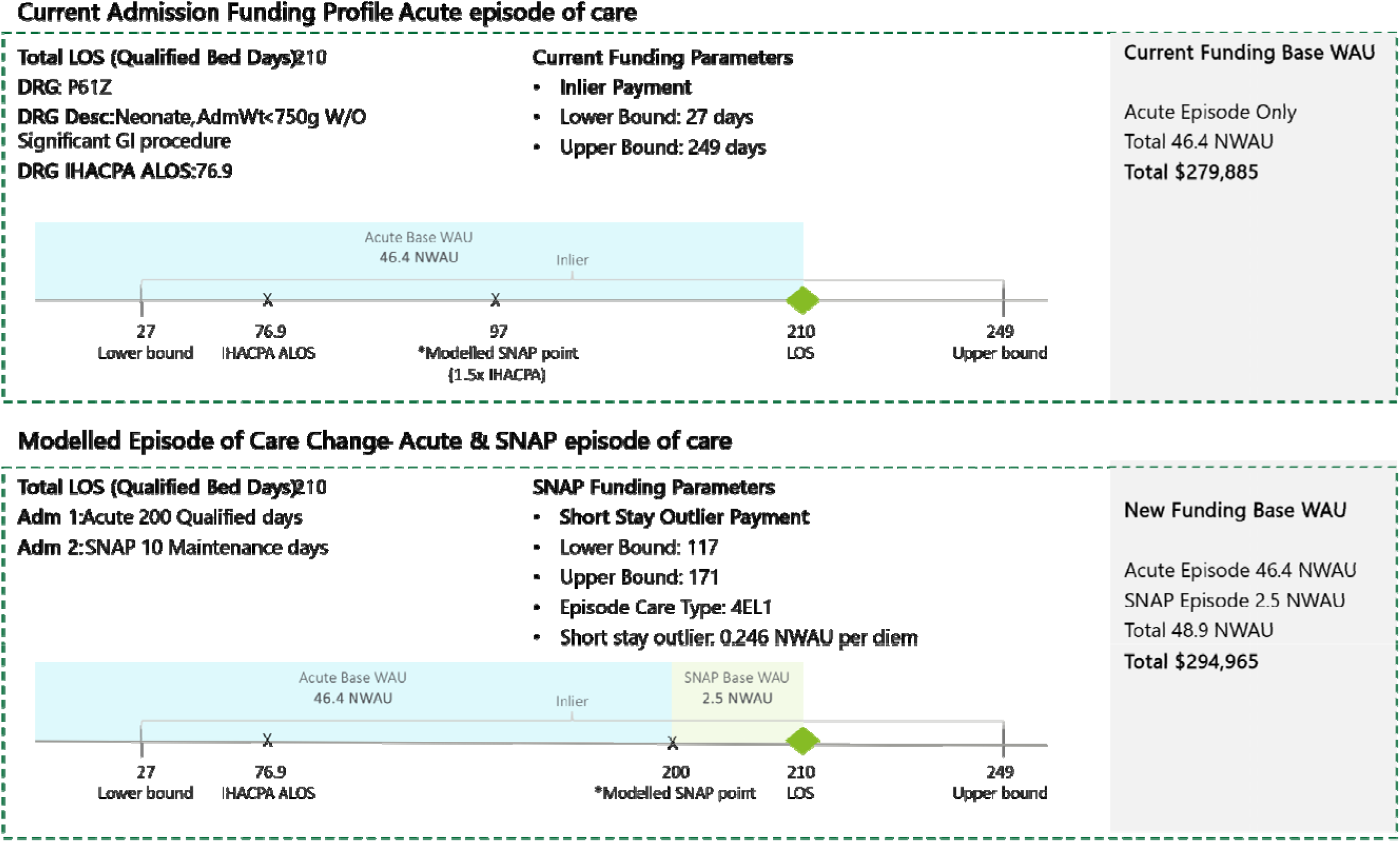
Case Study – Long-Stay Neonatal Admission – Lower Estimate. Independent Health and Aged Care Pricing Authority (IHACPA), Average Length of Stay (ALOS), Length of Stay (LOS), Weighted Activity Unit (WAU), Diagnosis-Related Group (DRG).

### Analysis

The lower estimate of the uplift was 92.3 NWAU (51 episodes) and the upper estimate was 485 NWAU (124 episodes), corresponding to AUD $556,616 and $2,926,011, in 2023-24 value. This is in the context of a modelled base for the activity of 2857 – 2895 NWAU ($17.2-17.5M). Long-term trends are visualised in Figure 6 and showed that this cohort is likely increasing in incidence within our centre.

**Figure 6.**
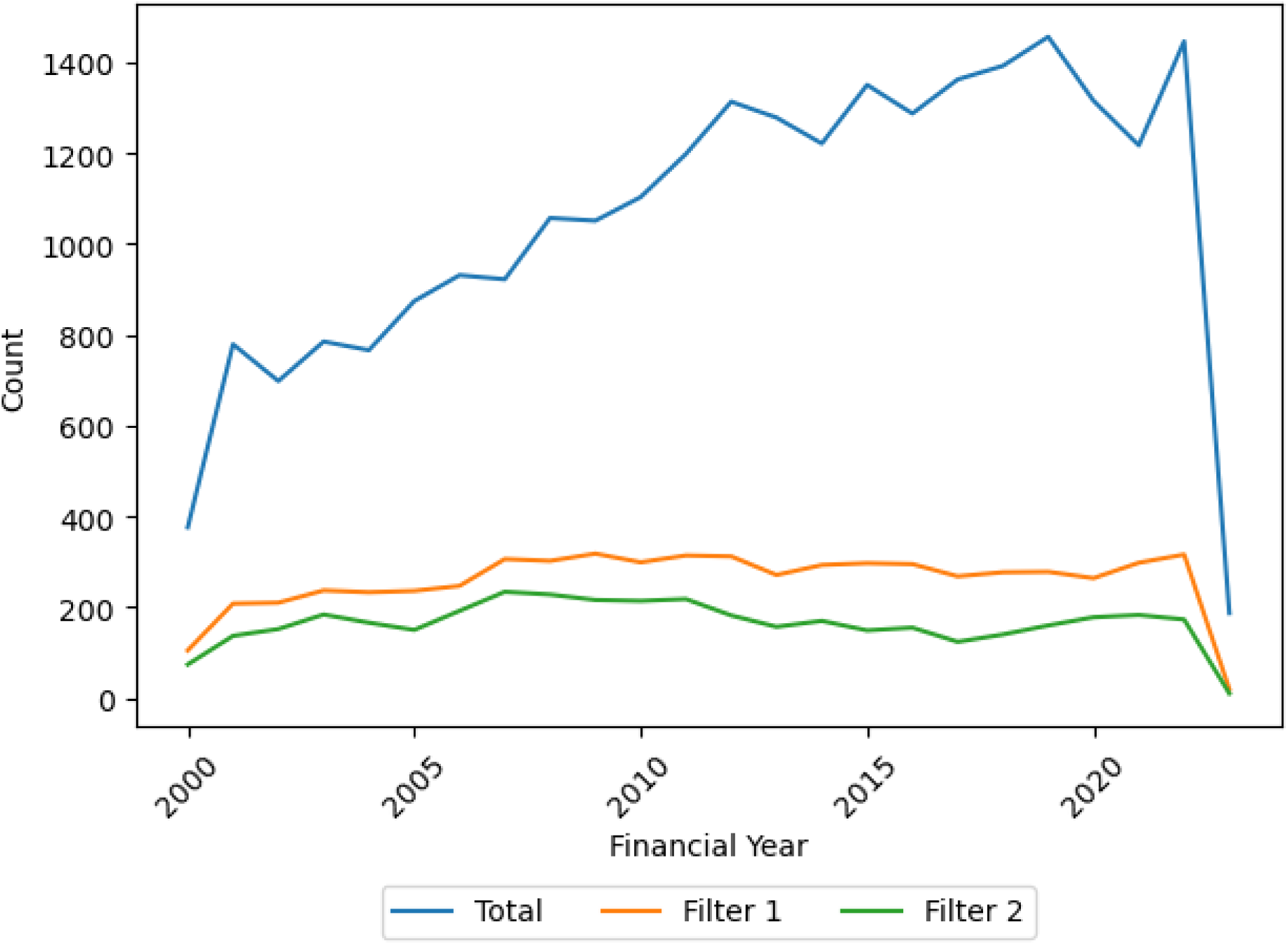
Long-Term Trends of SNAP-able Neonatal Admissions (2000-23)

## Discussion

### Estimated Financial Impact of Introducing SNAP Care

In this study of a level 6 neonatal unit, we identified between $0.5M and 3M in potential financial uplift from within the 2021-22 financial year, through utilising a care type change from acute admitted to SNAP. This is in the context of a base activity of $17.2-17.5M. When compared with the actual revenue from 2021 of $19.6M against a budget variance of $1.4M unfavourable, the potential uplift may be enough to account for the cost variance. With reference to long-term trends, this estimate appears to be at least stable, if not growing in potential impact.

### The Disconnect Between Funding and Clinical Practice

The challenges in aligning the NEP to good clinical practice are well described in submissions to IHACPA in recent years. Children’s Healthcare Australia describe scenarios in which bundled payments (where newborn care is included in the maternity care price), has significant limitations, including post-natal maternity mental health admissions. The issue of bundling ICU payments into neonatal DRGs is also highlighted, with the key message being that wide variability in length of stay, complexity, variability in the need for ICU care and variability in the type of ICU care utilised (Paediatric compared with Neonatal Intensive Care Units), make bundling less suitable in this context because the variance isn’t primarily accounted through an efficiency lens ^10^.

These challenges have led Queensland to unbundle neonatal from maternity funding ^13^. Within the feedback provided about this matter over multiple consultation periods, stakeholders noted that bundled payment does not reflect the current cost of care, that this could drive adverse resource allocation and that it may have the effect of separating mother and baby, which does not reflect current best practice ^14^.

### Challenges of Birth Weight-Based Funding Models

The Australia and New Zealand Neonatal Network submitted to IHACPA to move towards funding based on gestational age, given that this is a better predictor of clinical course than weight-based DRGs ^15^. Since DRGs (in version 12 and below) are based on data obtained from the National Minimum Dataset, and that dataset doesn’t currently collect information on gestational age, this is a key barrier to progressing this. Since the International Classification of Diseases Tenth Version (ICD-10-AM) Twelfth Edition has included greater detail around gestational age and this forms the basis for AR-DRGs, costed data is said not to be available to IHACPA until the AR-DRG 13.0 development cycle ^15^.

### Aligning Funding with Clinical Phases of Care

The academic and grey literature discussing neonatal care converges on a number of concepts-1) neonatal care is effective, costly and specialized; 2) neonatal care is provided in a variety of ways for a number of reasons, some of which include integration with the local health system and patient-specific needs (e.g. use of a Paediatric ICU rather than NICU for extra-corporeal membranous oxygenation in some centres); 3) There are likely to be three broad phases of care-hyperacute (e.g. intensive care), acute (e.g. special-care baby unit), sub-acute/maintenance/transitional care. Whilst ANZNN have submitted that a gestationally-orientated funding system will greater align with the clinical course of care, this doesn’t account for the system aspects of care pointed to in the SA context- that the duration of care intersects with the duration of each phase and is highlighted when each phase is routinely provided within a centre that provides all three phases of care, in contrast to the tacit separation that occurs when the phases of care are provided separately.

### The Economic Burden and the Cost of Neonatal Care in Context

The health economic and financing context of neonatal units, particularly in high income countries, reflects the complexities of providing high-quality, specialized care within the constraints of healthcare budgets and policies. Neonatal care, especially in NICUs, is resource-intensive, involving advanced technologies, highly specialized staff, and extensive use of pharmaceuticals and medical supplies ^16^. The cost of neonatal care can be substantial, with expenses for preterm or critically ill infants significantly higher than for term infants without complications. Future advancements such as critical care genomics or genomic newborn screening are likely to add further cost and clinical and operational complexities, leading to improved health outcomes ^17^.

However, the cost of the outcomes of newborn conditions such as prematurity undermanaged, are also substantial ^18^. Economic evaluations of interventions to prevent or treat newborn conditions are generally considered highly cost effective ^16^, ^19^. The benefits include significantly improved survival rates for extremely preterm and critically ill infants, reduced long-term disability, and enhanced quality of life for survivors ^20^. The investment in neonatal care not only saves lives but also contributes to reduced future healthcare costs, and improved developmental outcomes, supporting the argument that the high initial costs are justified by the substantial long-term benefits to individuals and society ^16^. This emphasizes the importance of continued investment in and optimization of neonatal care to ensure its sustainability and accessibility, aligning with broader health policy objectives of maximising health outcomes within available resources.

### Limitations of the Simulation Study

This exploration into care-type changes demonstrates a proactive approach to leveraging existing components of the ABF model (AN-SNAP) to enhance the financial sustainability of neonatal care and enable appropriate care. The limitations of the study are specific to the method used, and then questions about the appropriateness of applying the funding model to this context.

The limitations of the simulation are primarily that it provides an indication of the possible financial uplift in implementing the SNAP care type. The actual uplift would depend on implementation in practice and determining clinically appropriate indicators for care type change. This latter piece could be developed but there are currently no standardised tools available to guide when neonatal care shifts between the aforementioned phases of care. Given that neonates are entirely dependent for activities of daily living, current assessment instruments such as the FIM and paediatric modifications, wouldn’t be particularly helpful.

### Implementation Challenges

In our context, there were three significant practical challenges identified-1) whether the level of effort required to implement care-type change in this cohort is likely to return significant enough of a return (in terms of sustainability and impact on clinical care) to warrant the investment; 2) the existing implementation of neonatal ABF in SA has several constraints over and above that required by the IHACPA NEP model; 3) That changes would impact multiple systems, including operational financing and in particular, the PAS and interdependent Electronic Medical Record (EMR). SA uses the Sunrise EMR and PAS provided by Alere, but these problems are not inherent to the system, but rather related to SA’s configuration of the PAS functions in the system.

### Further Implications for Practice

Finally, the study serves as a reminder of the evolving nature of healthcare financing and the need for continuous review and adaptation of funding models to ensure they meet the changing needs of specialized care units. This project not only contributes to the body of knowledge on healthcare financing and neonatal care but also provides a practical framework for other units to follow, potentially leading to better healthcare outcomes and financial efficiency.

## Conclusion

In this study we simulated the financial uplift involved in introducing a care type change into neonatal care in our level 6 unit. This demonstrated a 2.8-17.4% uplift in funding. Exploring episode characteristics over 23 years demonstrated a long-term trend within our unit to an increase in this SNAP-eligible cohort. Understanding this further could be subject to future research.

We explored that there are several potential barriers to implementation, including practical steps to introduce this into the South Australian funding system and IT systems, as well as a lack of standardised tools available for determining when it would be clinically appropriate to change care type.

## Acknowledgements

Kirsty Taylor and Cameron Abbott from Deloitte Financial Advisory Pty. Ltd. provided advice and undertook the initial analysis.

## Author Contributions

DM is responsible for the concept, additional analysis and the manuscript.

## Data Availability Statement

Data is not available for sharing as this was not included as a part of the institutional approval.

